# Strategic use of SARS-CoV-2 wastewater concentration data could enhance, but not replace, high-resolution community prevalence survey programmes

**DOI:** 10.1101/2023.08.17.23293589

**Authors:** Cathal Mills, Marc Chadeau-Hyam, Paul Elliott, Christl A. Donnelly

## Abstract

Wastewater-based epidemiology (WBE) has been proposed as a tool for public health authorities to monitor community transmission of SARS-CoV-2 and other agents. Here, we review the utility of WBE for estimating SARS-CoV-2 prevalence using wastewater data from the Environmental Monitoring for Health Protection (EMHP) programme and prevalence data from the REal-time Assessment of Community Transmission-1 (REACT-1) study in England. Our analysis shows a temporally evolving relationship between wastewater and prevalence which limits the utility of WBE for estimating SARS-CoV-2 prevalence in high spatial resolution without a concurrent prevalence survey. We further characterise WBE for SARS-CoV-2 prevalence as i) vaccination-coverage-dependent and ii) variant-specific. Our work provides a gesopatial framework to map wastewater concentrations to public health boundaries, enabling public health authorities to interpret the relationship between wastewater and prevalence. We demonstrate that WBE can improve the cost efficiency and accuracy of community prevalence surveys which on their own may have incomplete geographic coverage or small sample sizes.

## Introduction

Wastewater-based epidemiology (WBE) was initially developed to monitor and investigate illicit drug use and the distribution of viruses. WBE involves collection of samples from wastewater treatment plants that capture human excretions through urine or faecal matter, and has enabled, for example, surveillance of poliovirus and 2009 influenza A (H1N1) [1, 2, 3]. During the COVID-19 pandemic, as governments and public health authorities around the world sought to monitor the transmission and spread of SARS-CoV-2 (the virus which causes COVID-19), WBE was applied as a disease surveillance tool, since SARS-CoV-2 is excreted in the faeces of both symptomatic and asymptomatic infected individuals. Clinically-confirmed SARS-CoV-2 prevalence estimates are sensitive to test-seeking biases, asymptomatic infections, and clinical testing capacity. Conversely, WBE represents an indirect, non-invasive, population-level disease surveillance tool for cost-effective, real-time monitoring of pathogen transmission [4, 5]. Several studies have established strong correlations between wastewater SARS-CoV-2 concentrations and reported COVID-19 cases [6, 7, 8], whilst others have estimated the time dependency and lead time (varying from four to ten days) from detection in wastewater to date of testing in clinical cases [9, 10, 11]. During the pandemic, SARS-CoV-2 prevalence in England was estimated for 45 sewage site catchments [12], whilst further research estimated the weekly viral wastewater concentrations in a spatially continuous domain [13].

Here, across 21 months of the COVID-19 pandemic in England, we provide a high-resolution spatiotemporal analysis and evidence synthesis of the utility of WBE for estimating SARS-CoV-2 prevalence. Our analysis uses data from one of the world’s largest community prevalence surveys; the REal-time Assessment of Community Transmission-1 (REACT-1) study and the Environmental Monitoring for Health Protection (EMHP) wastewater surveillance programme. Our geospatial framework maps wastewater concentrations from a sewage treatment plant to the level of a Lower Tier Local Authority (LTLA). Then, we i) quantify the relationship between wastewater concentrations and estimated infection prevalence from REACT-1, and ii) perform a modelling analysis to investigate the extent to which WBE can facilitate estimation of community prevalence of SARS-CoV-2.

## Results

### Relationship Between SARS-CoV-2 RNA Wastewater Concentrations and Prevalence

Consistent with our underlying scientific premise, in the *early* period (REACT-1 rounds 3 to 11, from 24 July 2020 to 3 May 2021), estimated wastewater concentrations were moderately-to-strongly correlated with SARS-CoV-2 prevalence (Spearman’s correlation *r* = 0.62; 95% CI: 0.59, 0.65), both in fine and coarse spatial resolutions (Supplementary Material Table SI 1). In the *late* studied period (rounds 12 to 19, from 20 May 2021 to 31 March 2022), we found the relationship between concentrations and prevalence to be complex, volatile, and temporally evolving. Within a subset of the five pre-Omicron rounds (rounds 12 to 16, 20 May 2021 to 14 December 2021) of the *late* period, we report weaker correlation (*r* = 0.28; 95% CI: 0.23, 0.33) between concentrations and prevalence.

We propose here use of the estimated prevalence per log concentration in wastewater to capture the prevalence-to-wastewater relationship, and hence the implied time-varying population-level faecal shedding. Relationship complexity appeared due to i) rapid rollout of heterogeneous vaccination of LTLA populations nationally, and ii) rapid replacement of the Delta variant by the Omicron BA.1 and BA.2 sub-variants. For rounds 12 to 19, the estimated prevalence per log concentration was moderately correlated (*r* = 0.58; 95% CI: 0.55, 0.61) with the estimated proportion of an LTLA population that was fully vaccinated (two or more doses of any vaccine), though correlation was stronger when we instead considered the population proportion fully vaccinated in the previous round (*r* = 0.66; 95% CI: 0.63, 0.68).

Thus, in estimating the association between SARS-CoV-2 prevalence and wastewater concentrations, it is important to account for the proportion of a population fully vaccinated and specifically the percentage of vaccinated individuals in the preceding month. The implied lag may reflect the time taken for a vaccine to become effective at reducing faecal shedding. Inclusion of the vaccination-log concentration interaction (defined in Supplementary Material Table SI 2) increased the wastewater-infection prevalence correlation (*r* = 0.71, 95% CI: 0.69, 0.73) relative to the unadjusted model.

By mid-December 2021, the REACT-1 study and EMHP wastewater programme had found that Omicron BA.1 had become the dominant SARS-CoV-2 variant in England [14, 15]. Here, we show that the percentage testing positive per log concentration in wastewater increased substantially during the Omicron period, indicating less faecal shedding at a population level (Figure 1)

**Figure 1.**
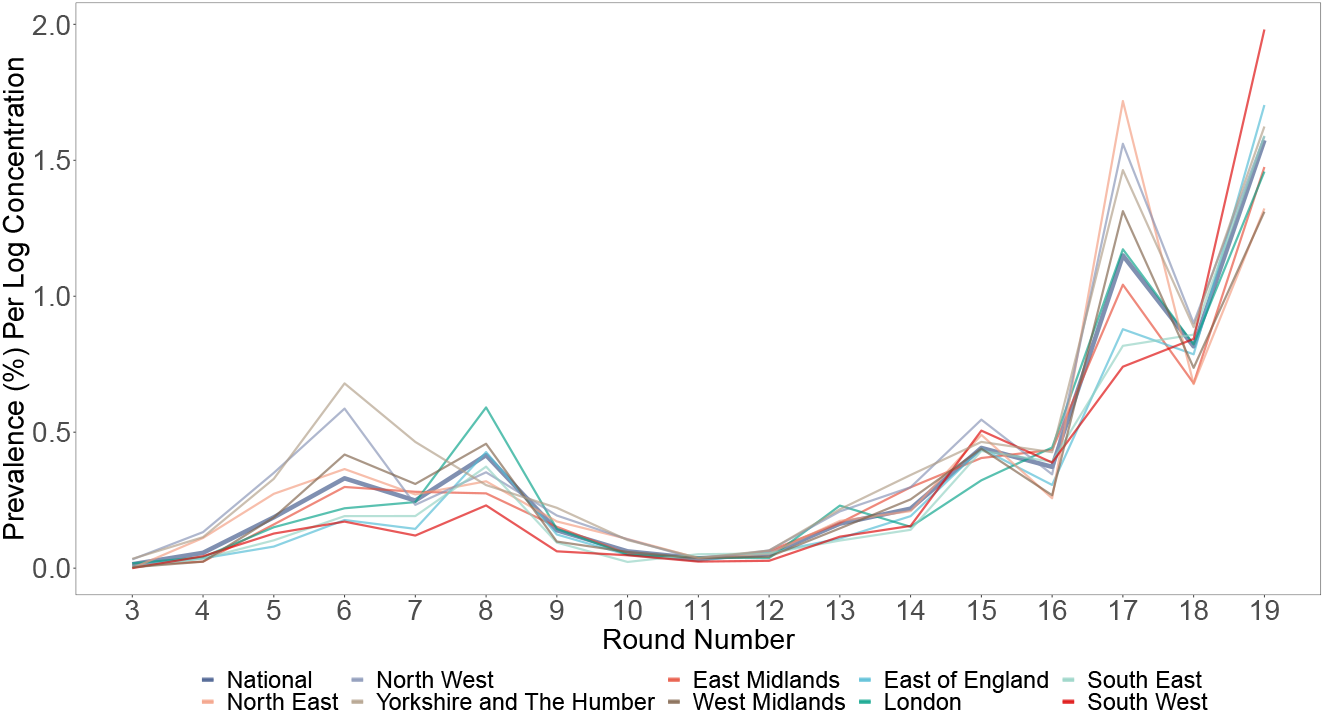
Prevalence-to-wastewater relationship. Weighted regional and national averages of the REACT-1 prevalence per estimated log concentration by round of data collection, from 24 July 2020 to 31 March 2022. Greater values of the ratio correspond to lower implied faecal shedding per positive individual. For reference, vaccination begins in round 8 (6 January 2021), full vaccination starts to become prevalent from round 12 (20 May 2021) onwards (when the survey round estimate of the national average proportion of the entire population fully vaccinated is 31.7%), the Delta variant became dominant between rounds 11 (15 April 2021) and 12, and the Omicron BA.1 and BA.2 sub-variants are dominant in rounds 17 to 19 (from 5 January 2022 to 31 March 2022).

### Out-of-Sample Wastewater-Model-Based Estimates of REACT-1 Prevalence

The *early* period of our modelling analysis (rounds 3-11, from 24 July 2020 to 3 May 2021) corresponded to a duration of relatively low, stable prevalence levels (mean LTLA estimate: 0.55%) and vaccination was at low levels (mean fully vaccinated proportion for an LTLA in round 11 was 12.2%). Conversely, the ten-month *late* period (rounds 12-19, 20 May 2021 to 31 March 2022) incorporated expanded wastewater testing coverage and the epidemic’s evolution was characterised by high prevalence levels, heterogeneous vaccination of the population, and occurrence of a new, highly transmissible Omicron variant with different properties, such as lower viral loads, selective reduction in Omicron infectivity in the intestinal tract, and shorter duration of respiratory shedding, compared to the Delta variant [16, 17, 18].

Employing an iteratively-updated wastewater-based model, out-of-sample prevalence estimates were generally noisy at an LTLA level in the *early* period (Supplementary Material Table SI 4). Failure to detect extreme prevalence peaks and sub-optimal accuracy for detection of prevalence level change argues against relying exclusively on a wastewater model post-calibration for high-resolution inference. We report more accurate prevalence estimates at the coarser spatial resolution of regional level, as displayed for the subset of rounds 9 to 11 (Figure 2). Thus, our data show that without a concurrent prevalence survey, policy-makers could attain a reliable representation of regional prevalence trends and whether they are experiencing increasing or decreasing prevalence levels. Likewise, we could reliably predict national prevalence estimates, and the direction of prevalence changes in each survey round were all correctly detected (Supplementary Material Figure SI 1).

**Figure 2.**
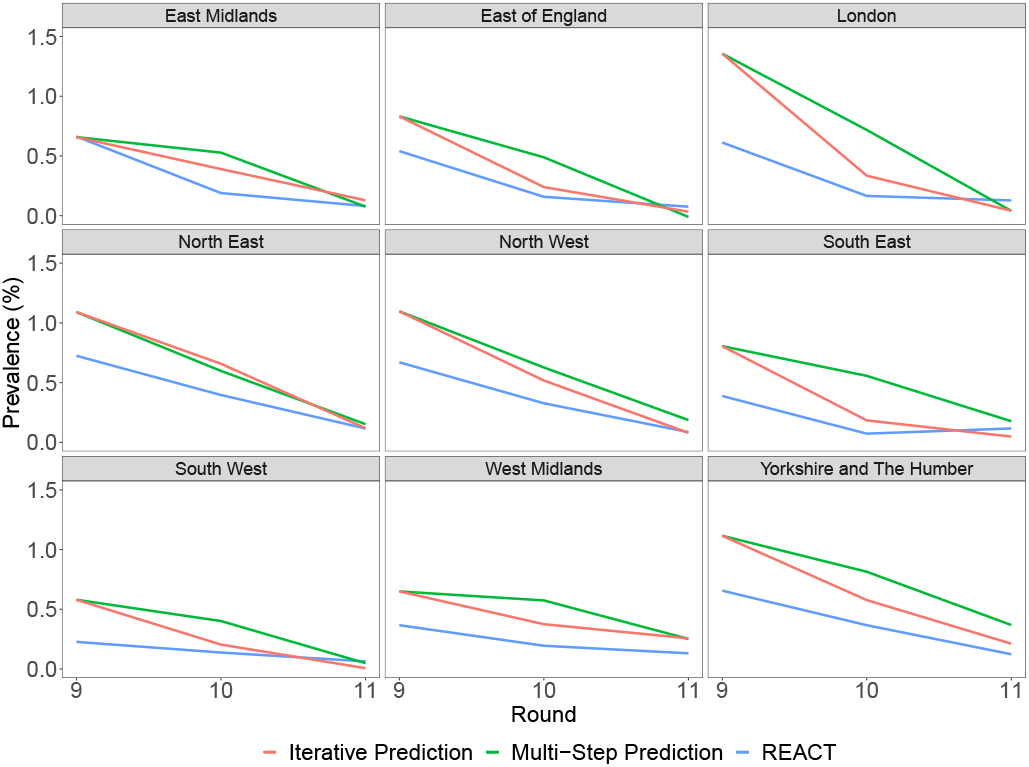
Regional out-of-sample prevalence estimates in rounds 9 to 11 (4 February 2021 to 3 May 2021). Wastewater-model-based prevalence estimates are shown alongside REACT-1 prevalence (blue). The model was trained at an LTLA level and out-of-sample regional estimates were obtained for rounds 9 to 11 i) with iterative updating of the model (red) and ii) without model recalibration (green). The two scenarios enable comparison of estimates with, and without, continuous model calibration. Regular over-prediction is a result of the decrease in estimated prevalence per log concentration around rounds 8 to 10 (6 January 2021 to 30 March 2021) indicative of increased faecal shedding), whilst the ratio stabilised between rounds 10 and 12 (11 March 2021 to 7 June 2021), as displayed in Figure 1.

However, for multi-step out-of-sample estimates (estimating out-of-sample prevalence in several individual time periods without model recalibration), the wastewater-based model did not provide an accurate high-resolution account of LTLA-level prevalence, although we again attained relatively strong predictive performance at a regional level (Figure 2 and Supplementary Material Table SI 3). This is despite a continued absence of any coincident prevalence survey, albeit with persistent over-prediction due to uncalibrated changes to the prevalence-to-wastewater relationship. Here, the region-level estimates derived by multi-step testing (Mean Absolute Error, MAE = 0.26%) were marginally inferior to those derived by regular, iterative updating of the prevalence-to-wastewater relationship noted above (MAE = 0.20%).

In the *late* period (20 May 2021 to 31 March 2022), irrespective of the modelling environment of either iterative updating or multi-step testing without model recalibration, the out-of-sample wastewater-model-based LTLA-level prevalence estimates remained sub-optimal, albeit detecting 86.0% of prevalence changes in the Omicron BA.2 peak (round 19) via the iteratively-updated model. Indeed, even if there were only a single model update, say round 17 (i.e. omitting REACT-1 round 18), during the Omicron wave of rounds 17 to 19 (from 5 January 2022 to 31 March 2022), the wastewater model enabled detection of 87.4% (95% CI: 83.2%, 90.9%) of the within-round 19 LTLA-level prevalence changes.

In coarser spatial resolutions, in contrast to the model for multi-step estimates, regional and national prevalence changes were well-detected by the iteratively-trained model, including 85.2% (95% CI: 66.2%, 95.8%) of out-of-sample (*n* = 27) regional prevalence changes (Figure 3). A caveat is that, for round 17, our trained wastewater model could not account for large initial prevalence surges induced by the Omicron variant. Reduced levels of detected population-level faecal shedding were apparent as wastewater concentrations (and the wastewater-vaccination interaction) did not show the same magnitude of increases as prevalence levels. Nevertheless, following further calibration in rounds 17 and 18, closer correspondence was observed by round 19 between out-of-sample wastewater-based regional estimates and REACT-1 prevalence levels (Figure 3).

**Figure 3.**
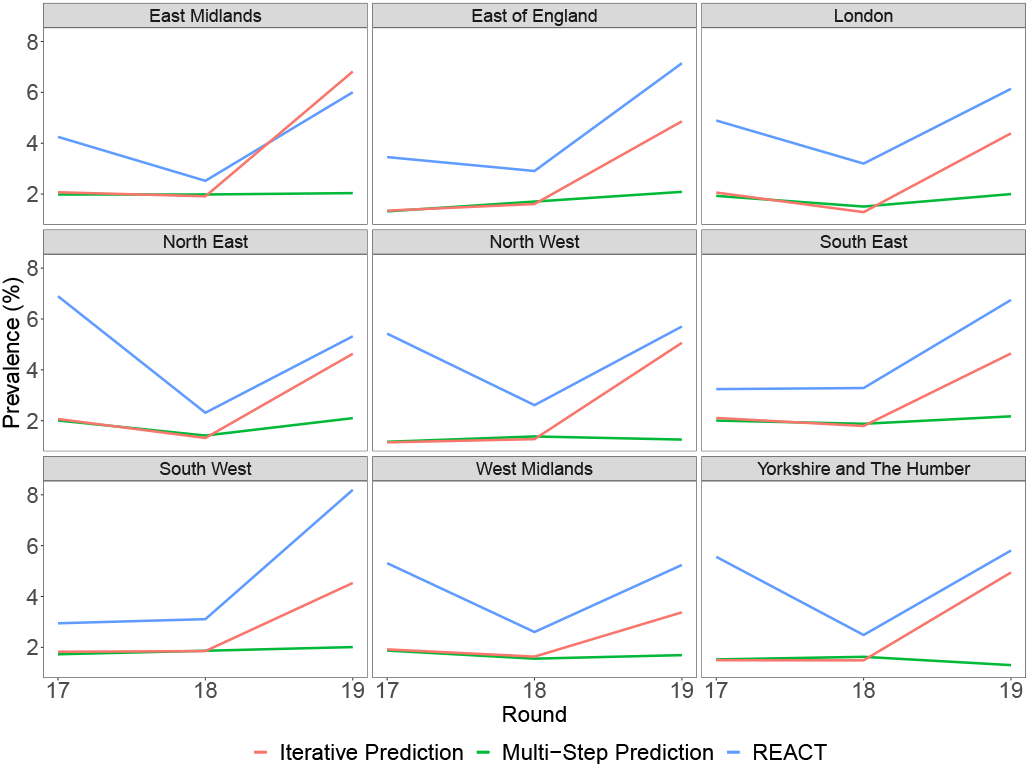
Regional out-of-sample prevalence estimates for the Omicron wave. Wastewater-model-based regional prevalence estimates are shown, alongside REACT-1 prevalence (blue) for rounds 17 to 19 (from 5 January 2022 to 31 March 2022). Similar to Figure 2, the model was trained at an LTLA-level and out-of-sample regional estimates were obtained for rounds 17 to 19 i) with iterative updating of the model (red) and ii) without model recalibration (green). The iteratively-updated model enabled out-of-sample prevalence estimates to adapt to the Omicron-induced changes to the prevalence-to-wastewater relationship yet persistent under-prediction was likely due to reduced population-level faecal shedding.

**Figure 4.**
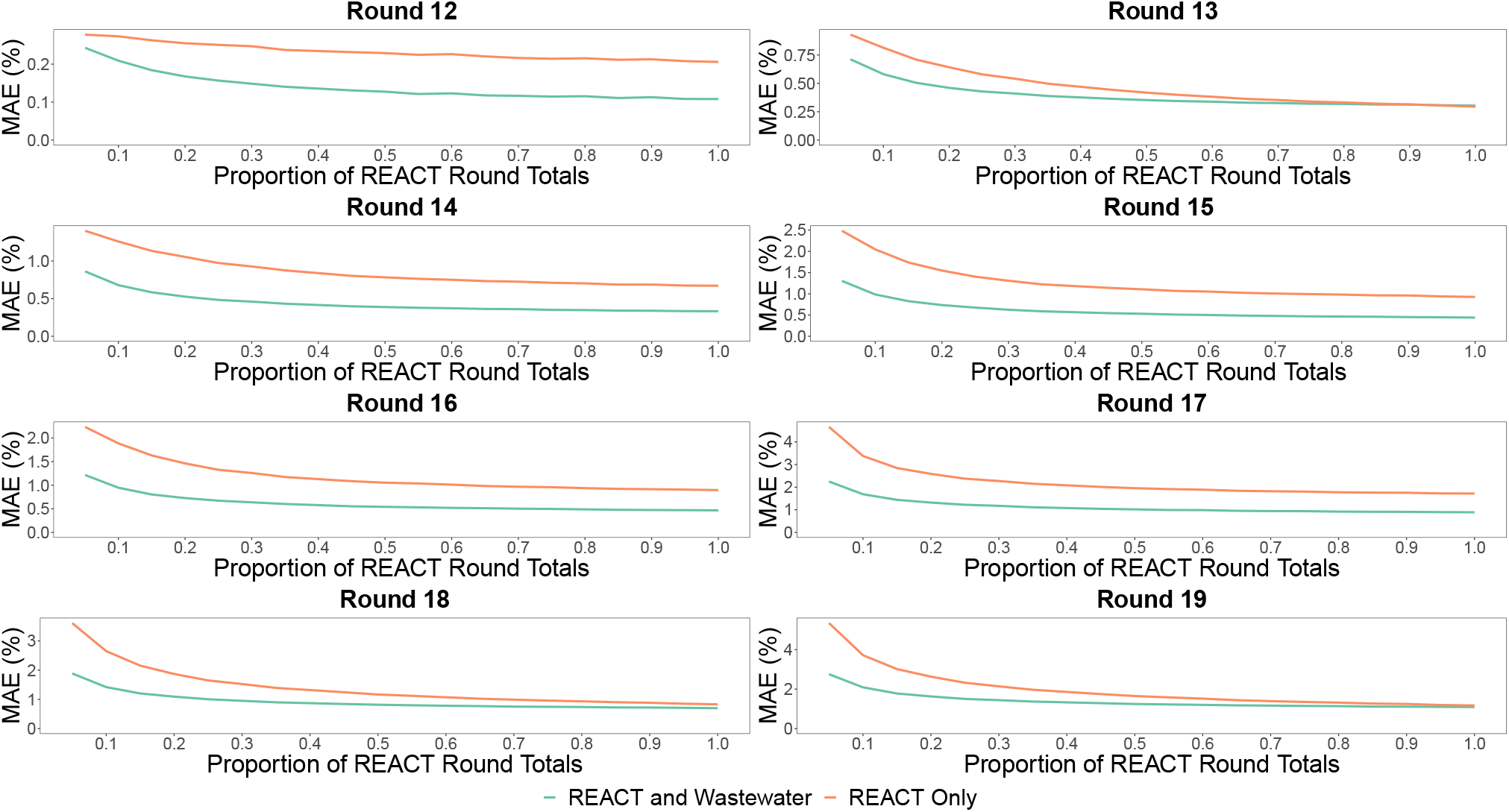
Prevalence estimate accuracy in simulations of reduced survey sample size with and without wastewater modelling. In individual rounds of 12 to 19 (from 20 May 2021 to 31 March 2022), the following procedure was replicated 100 times for proportions ranging from 5% to 100% in intervals of 5%: Simulated environments included 5% to 100% of each LTLA’s REACT-1 survey round sample size totals and the number of positives was simulated binomially from the reduced survey total with success probability equal to the REACT-1 weighted prevalence. Then, for each proportion, reduced-sized-survey-based prevalence levels were used to calibrate wastewater-model-based estimates. Both the reduced-survey-based prevalence levels (orange) and combined reduce survey-wastewater estimates (green) were compared to spatially smoothed REACT-1 prevalence levels via the Mean Absolute Error (MAE) within each of the 100 simulation replicates, and the mean average MAE is visualised above.

### Complementary Use of WBE for Monitoring SARS-CoV-2 Prevalence

We have shown sub-optimal wastewater-based model performance in out-of-sample scenarios without concurrent prevalence surveys due to the temporal inconsistency of the prevalence-to-wastewater relationship. We now assess utility of WBE to complement and enhance prevalence surveys in representative, high-resolution estimation of SARS-CoV-2 prevalence for both economic and logistical efficiency.

Results are presented for varying (counterfactual) intensities of community prevalence surveys. To ensure robustness of inferences, we focus primarily on results for the *late* period (rounds 12-19, 20 May 2021 to 31 March 2022) due to more complete, representative wastewater surveillance. Our approach here is two-fold as we examine i) environments with reduced geographic coverage, and ii) environments with reduced survey round sample size.

First, for the scenario with varying LTLA-level survey coverage, across the varying proportions of LTLAs within each round that were used for model training, to enable equitable comparisons, we specified a random 10% of withheld LTLAs from each survey round for wastewater-model-based prevalence estimation. Fixed test set sizes within each fold enabled equitable comparisons of model performance. We replicated the procedure 50 times, across different training-testing environments of the same proportions, to reduce sensitivity of inferences to individual training and testing sets.

As expected, relative to REACT-1 prevalence levels, maintaining a higher geographic coverage of prevalence surveys throughout the ten-month period produced more accurate out-of-sample wastewater-based prevalence estimates (Table 1 and Supplementary Material Figure SI 2). Nevertheless, training environments with fewer LTLA-level observations enabled representative indication of rising or declining prevalence levels for the omitted LTLAs throughout the sustained period, although the precise estimates tended to depart from REACT-1 prevalence levels. Similar results were attained for training-testing environments in the *early* period of 24 July 2020 to 3 May 2021 (Supplementary Material Table SI 5), as higher training survey coverage yielded more accurate estimates of test set prevalence, although increasing coverage from 80% to 90% of a REACT-1 round’s LTLAs yielded marginally inferior predictive performance (possibly due to low LTLA-level wastewater coverage). The representative out-of-sample prevalence estimates for omitted LTLAs, in the presence of a concurrent, reduced-scale prevalence survey, is an apparent consequence of the spatial consistency of the prevalence-to-wastewater relationship (Supplementary Material Figure SI 3), particularly in the *late* period with expanded wastewater surveillance where estimated population-level faecal shedding rates became increasingly consistent across LTLAs.

**Table 1:**
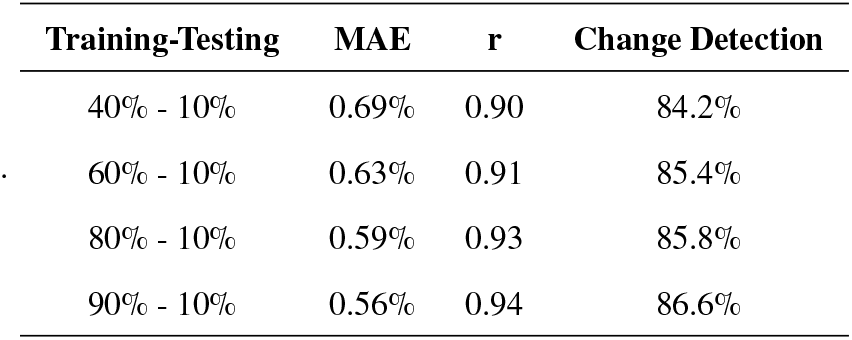
Wastewater-based complementary prevalence estimates across rounds 12 to 19 (20 May 2021 to 31 March 2022) using varying geographic coverage of prevalence surveys for training. Wastewater-based models were trained using 40% to 90% of each REACT-1 round’s observations and 10% of observations per round were used for real-time, out-of-sample wastewater-based prevalence estimation. Identical test sets of equal size were used within each fold to enable comparison of the model-based information offered by various training environments. The average test set prevalence across the 50 folds was 2.25%. MAE represents the median of Mean Absolute Error values and *r* represents the median Pearson’s correlation coefficient.

Second, we assessed how wastewater surveillance could enable reductions in the number of participants within survey rounds and thus contribute to greater cost efficiencies in prevalence surveys. Across each of the varying reduced survey round totals, relative to the spatially-smoothed prevalence levels (see Materials and Methods), we observed a consistent trend of wastewater-model-based estimates improving upon the corresponding reduced-survey-based prevalence estimates. Within each round, the average MAE expectedly declined as the survey round sample size totals increased for both the reduced-survey-only and combined survey-wastewater environments, although the wastewater model consistently improved upon the reduced survey for almost every round and survey sample size total. Indeed, irrespective of the choice of smoothing method (Kriging model or kernel smoothing) and whether we attempted to spatially smooth prevalence estimates from reduced survey totals (with and without wastewater), wastewater-based modelling improved the accuracy of prevalence estimates from reduced surveys.

## Discussion

Our spatiotemporal analysis provides a detailed evidence synthesis of the utlity of WBE. We identify that WBE alone is insufficient for high-resolution monitoring of SARS-CoV-2 prevalence without concurrent prevalence surveys, whilst reliable wastewater-based prevalence monitoring at a coarse spatial resolution is contingent on recent calibrating community prevalence surveys. However, wastewater-based modelling can play an important complementary role in improving the cost efficiency of prevalence surveys by filling gaps in spatial coverage of prevalence surveys or by enhancing accuracy of reduced-size surveys. Therefore, appropriateness of WBE is dependent on the use case (WBE alone or complementary to prevalence surveys), spatial resolution (coarse or fine), and concurrent epidemic dynamics (such as vaccination and variants).

Looking at the ratio of SARS-CoV-2 prevalence in the population to (log-transformed) viral particle concentration in wastewater reinforced the idea that there are two key factors at work in the temporally-evolving prevalence-to-wastewater relationship; i) levels of vaccination coverage and ii) differences in faecal shedding according to predominant variants. Reduced population-level faecal shedding in the aftermath of widespread vaccination is consistent with findings from community-based wastewater surveillance of COVID-19 in educational facilities [19]. Furthermore, variant-specific faecal shedding rates (specifically lower shedding rates during the Omicron period) may have led to breakdowns in the relationship between wastewater concentrations and prevalence. However, the additional breakdowns in the relationship at the beginning of the Omicron wave cannot be disentangled from the greater background vaccination and immunity, compared to previous waves. Indeed, changes in the relationship had already been observed in the preceding months (from September 2021), possibly due to changes to the dominant SARS-CoV-2 variant (as the Delta variant became dominant in the English population) [14]. Nevertheless, similar indications of reduced faecal shedding during the Omicron wave were observed in a community-based analysis of wastewater in the United States [20]. While our findings are at the population (rather than individual) level, they are consistent with clinical data for the Omicron variant which indicate preferential infection of the upper airway, selective reduction in Omicron infectivity in the intestinal tract, as well as lower viral loads and shorter duration of respiratory shedding, compared to the Delta variant [16, 17, 18, 21].

Across both periods of our analysis, a trained wastewater-based model alone did not provide accurate, high-resolution prevalence estimates in settings without a concurrent prevalence survey. In the *early* period (24 July 2020 to 3 May 2021), deviating prevalence estimates may be due to noisy LTLA-level estimated wastewater concentrations due to the smaller number of sampled neighbouring LTLAs (in these rounds), and/or the changing prevalence-to-wastewater relationship (as vaccination commenced and SARS-CoV-2 variants evolved). In the *late* period (20 May 2021 to 31 March 2022), weak out-of-sample predictive performance appears to be a direct consequence of the temporally unstable relationship between wastewater and prevalence (apparently due to both vaccination and the emergence of the Omicron variant). Our analysis identified a lack of temporal consistency that is compatible with the findings of [11], where the wastewater-to-clinical cases ratio also changed substantially during the pandemic. Limitations of our analysis include the relatively sparse geographical coverage of wastewater collection sites in England (especially earlier in the pandemic), sensitivity of the wastewater surveillance to quantify RNA concentrations [22], and also imprecision of the REACT-1 prevalence survey data at the LTLA level, as well as possible inaccuracies in the weighted estimates employed to correct for variable response rates in different sectors of the population [23]. Further limitations (discussed in Supplementary Material 6 and Supplementary Material 7) include the temporal resolution of our data and discrete time periods of data collection, alongside the lack of adjustment for variability in estimated wastewater concentrations and the averaging of concentrations over a survey round and across treatment plants.

Unlike high-resolution settings, in coarser spatial resolutions, our wastewater-based model generally provided a reasonable estimate of out-of-sample national and regional trends for up to three months without a simultaneous prevalence survey, albeit dependent on concurrent epidemic dynamics and the prevalence-to-wastewater relationship. In particular, multi-step estimates of regional prevalence without any model recalibration were not representative of sustained prevalence surges during the Omicron wave. Indeed, a single additional round of model recalibration during the Omicron wave enabled more accurate prevalence estimates. Hence, we confirm the importance of prevalence surveys for estimation and model calibration, alongside a potential use case of WBE; this could possibly be undertaken between rounds of a prevalence survey to improve continuity of regional and national prevalence estimates, contingent on availability of vaccination and variant tracking to inform the updating of the prevalence-to-wastewater relationship.

Importantly, by leveraging the spatial consistency of the prevalence-to-wastewater relationship (Figure SI 3), we show that wastewater-based-modelling can improve logistical efficiency of prevalence surveys by providing prevalence estimates over sustained periods for geographies not covered by prevalence surveys. Similarly, relative to spatially-smoothed prevalence, for simulations of reduced survey round sample size, combined survey-wastewater-based estimates consistently improved upon the accuracy of reduced survey prevalence levels alone, even for extremely small-sized surveys. Therefore, we demonstrate that combining WBE with intermittent survey data or fewer samples of survey data may help to maximise the geospatial reach, accuracy, and value of the information, while reducing the cost. Evidently, WBE data could be used to provide additional, reliable information to “fill in the gaps” in estimating SARS-CoV-2 prevalence during intervals with reduced survey data. This could be a cost-effective approach, since the WBE data are relatively cheap to obtain compared to conducting population surveys. Further work including an economic evaluation would be needed to model different scenarios based on timing, extent, and duration of survey data and geospatial coverage of the wastewater data, but this is beyond the scope of the current work. For example, a cost-benefit analysis for Germany estimated that the national cost for WBE surveillance reagents would be only 0.014% of those required for clinical testing [24], although this did not account for the relatively unknown value (accuracy) of information from WBE compared to prevalence survey data.

From a public health policy perspective, our analysis has shown that WBE alone is insufficient for modelling SARS-CoV-2 prevalence at high (spatial, temporal) resolution, although it may provide useful information on trends at a national or regional level and also complement prevalence surveys to achieve greater cost efficiencies via reduced-sized surveys. In the event of future outbreaks of infectious diseases, a combination of surveillance by survey data and WBE is likely to be the most cost-effective approach to obtaining situational awareness for policy makers. This will require concomitant monitoring of vaccination coverage and variants (accessible via prevalence surveys, WBE, and genotyping), alongside clinical indicators of epidemic dynamics. Together, these will help to determine the optimal timing of wastewater model recalibration and the scale of survey and wastewater monitoring required to ensure representative prevalence estimation.

## Materials and Methods

### Environmental Monitoring for Health Protection (EMHP) Programme

Wastewater concentration data were sourced from the EMHP programme, which was led by the United Kingdom Health Security Agency (UKHSA) and tested untreated sewage across England for fragments of SARS-CoV-2. The objectives of the programme were to monitor wastewater viral concentrations of SARS-CoV-2 RNA, variants of concern, and variants under investigation. For estimation of wastewater concentrations, samples were collected three to four times weekly at sewage treatment works (STWs) and sewer network sites [14].

The monitoring programme commenced in July 2020 at 45 STWs and a large-scale expansion occurred nationally in June 2021. There are two ways that wastewater samples are obtained: either via i) grab samples (a single sample taken using a small container at one point in the day), or ii) composite samples (an autosampler samples at regular intervals throughout the day and mixes the samples together in a container). See Supplementary Material 7 for discussion alongside further sources of wastewater uncertainty including sample volume, temperature and time-induced decay, inherent wastewater variability (possibly due to precipitation and dilution effects), and the Theoretical Limit of Detection (TLoD) [14, 22].

Viral concentrations of SARS-CoV-2 RNA were obtained from wastewater samples by quantifying the number of SARS-CoV-2 N1 gene copies per litre via the Reverse Transcriptase Polymerase Chain Reaction (RT-qPCR) process, a quantification method which combines two main steps of reverse transcription (RT) and quantitative PCR (qPCR) [7]. Normalisation of wastewater flows is necessary to account for precipitation dilution of concentrations of SARS-CoV-2 RNA. The indirect normalisation conducted under the EMHP programme is described elsewhere [22], but briefly, the technique assumed that wastewater flows were not directly observable, flow variability was estimated and outliers were identified. The final output was the flow-normalised viral concentrations of SARS-CoV-2 RNA, defined in terms of numbers of gene copies per litre (henceforth termed the *concentrations*), which are then log-transformed for our analysis.

### REal-time Assessment of Community Transmission-1 (REACT-1) Study

Community prevalence data were sourced from the REACT-1 study which obtained estimates of prevalence of SARS-CoV-2 infection in England from 1 May 2020 to 31 March 2022. Across 19 distinct rounds of cross-sectional surveys [25], random samples of the English population (over 5 years of age) were taken [26]. Rounds of REACT-1 were approximately monthly with durations between 15 and 31 days.

The REACT-1 study involved participants using a self-administered throat-and-nose swab kit and completing a questionnaire. Throughout the 19 rounds, approximately 2.39 million respondents were tested for SARS-CoV-2 by RT-PCR, of whom approximately 25,300 individuals tested positive. An advantage of the use of random samples was that it avoided the inherent biases that exist with only testing of symptomatic individuals, test-seeking behaviours, and the availability of tests. In addition to identifying spatiotemporal trends in prevalence, the study enabled analysis of variants, vaccine effectiveness, and vaccine coverage, all of which became increasingly important as the COVID-19 pandemic progressed [26, 27].

### Geospatial Mapping of Wastewater Concentrations and Relating to REACT-1

The catchment areas of STW locations for the reported flow-normalised concentrations do not align with the spatial resolution of the REACT-1 study; the 315 LTLAs of England. We therefore used a geospatial approach to map the wastewater concentrations from the STW level to LTLA level. We employed the lookup table provided by [28], which gives a mapping of wastewater catchment areas to Lower layer Super Output Areas (LSOAs) which are small regional geographies that combine to form the larger geography of an LTLA. For 21 STWs of the EMHP programme which are not included in the lookup table, to estimate their catchment areas, we employed an approximation method proposed by [13].

Equipped with the catchment-to-LSOA mapping area data and Office for National Statistics (ONS) 2019 mid-year population estimates, we derived geospatial population estimates (GPEs) for the intersection area between the catchment area of STW *i* and LTLA *k*. Our geospatial approach is adapted from [28], and we assumed that each LSOA population is uniformly distributed across the area serviced by STWs:

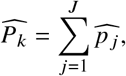

where *P*_*k*_ is the GPE for LTLA *k* and 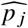 is our estimate of the serviced population of LSOA *j* based on our population estimates for the intersection area between the catchment area of STW *i* and LTLA *k* (See Supplementary Material 6 for further details).

For each LTLA *k*, the weight *w*_*ik*_ assigned to STW *i* for concentrations was the ratio of the GPE for the intersection area between the catchment area of STW *i* and area of LTLA *k* divided by the GPE for LTLA *k*:

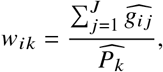

where *g*_*ij*_ are the GPEs for the intersection area between the catchment area of STW *i* and area of LSOA *j*. If any particular STW *s* failed to report concentrations within a round of the REACT-1 study, the weights of LTLA *k* for that round were re-weighted such that ∑_*i*≠*s*_ *w*_*ik*_ = 1 and *w*_*sk*_ = 0.

For each STW *i*, we took the median concentration across the REACT-1 round. Then, the estimated concentration of an LTLA for a given round was the weighted average of the concentrations of STWs within that LTLA. The weights employed here were the STW-LTLA population estimates. A sensitivity analysis (Supplementary Material 8) considered alternative approaches such as taking the wastewater measurement of the (single) dominant STW in an LTLA, the mean concentration across the round, and various lead/lag times (where we expanded/reduced the window for wastewater measurements). Each alternative approach led to a diminished relationship between the wastewater concentrations and REACT-1 prevalence levels.

In the event of future outbreaks of infectious diseases, the developed approach could be applicable to population-level wastewater analyses which require alignment of wastewater catchments to geographies used by public health authorities. Nevertheless, there exists limitations with the predictive approach of using GPEs, such as transient populations, uncertainty surrounding population estimates of small geographic areas, and potentially unrepresentative wastewater concentration estimates for some LTLAs, each of which are discussed in (Supplementary Material 6).

### Modelling REACT-1 SARS-CoV-2 Prevalence

Our spatiotemporal analysis is divided into two distinct periods; the *early* period of rounds 3 to 11 (24 July 2020 to 3 May 2021 - the first time periods where the EMHP programme intersects with the REACT-1 study) and the *late* period of rounds 12 to 19 (20 May 2021 to 31 March 2022). The two periods are analysed independently due to large differences in wastewater surveillance coverage and the EMHP programme’s transition from usage of two laboratories to a single laboratory.

Our primary model for estimating LTLA-level SARS-CoV-2 prevalence is an extreme gradient boosting model which is implemented via the xgboost package in R version 4.2.1 [29, 30]. Briefly, the extreme gradient boosting (known as XGBoost) machine learning algorithm is comprised of an ensemble of weak learners consecutively fit to data in a greedy manner. The algorithm is a parallelised, optimised version of the general gradient boosting algorithm [31]. XGBoost was chosen due to its predictive capabilities and highly flexible nature (ability to handle non-linearities and time-varying relationships), whilst also enabling regularisation (via model hyperparameters such as the learning rate) and cross-validation. Despite the discriminative nature of the gradient boosting approach, we retained the ability to appraise variable importance for predictors of prevalence in our training environments. We developed a Bayesian hierarchical model to ensure robustness of conclusions from our modelling analysis (described in Supplementary Material 9). Covariates used in our models are outlined in Table SI 2, and are wastewater-based variables with the exception of our proposed log concentration-vaccination interaction for the *late* period (discussed in Supplementary Material 10).

### Types of Wastewater-Model-Based Estimates

Our modelling analysis focused on out-of-sample wastewater-model-based estimates of LTLA-level prevalence. Regional and national estimates were achieved by weighting the LTLA-level estimates by corresponding LTLA populations.

There were three broad types of out-of-sample estimates: i) *Iterative training and testing* involved calibrating our wastewater model using a minimum of four REACT-1 rounds and subsequently using wastewater data to estimate prevalence for a full REACT-1 round one-at-a-time. The idea was to evaluate inferences gained from subsequently relying on WBE alone or temporarily between rounds of a prevalence survey. ii) *Multi-step testing* similarly involved training our wastewater model for five to six rounds but using wastewater data to estimate prevalence for three individual rounds without recalibrating the model. The intuition here was to appraise the exclusive application of WBE over an approximate three-month period without a concurrent REACT-1 survey to recalibrate the prevalence-to-wastewater relationship. iii) *Complementary wastewater-based estimation* involved counterfactual incomplete survey rounds, where we assessed the effectiveness of WBE for complementing incomplete prevalence surveys which could arise due to funding and/or logistical limitations which prevent a large-scale, national survey programme. First, we considered reduced geographic coverage of prevalence surveys as we excluded varying numbers of random LTLAs from single survey rounds, and the corresponding prevalence levels were estimated using our wastewater-based-model and compared to corresponding REACT-1 prevalence estimates. Second, we investigated how WBE could contribute to prevalence estimation in the event of reduced survey sample size totals within rounds. Using reduced survey round totals, we simulated the corresponding number of positive individuals (in a round) from a binomial distribution with success probability equal to the REACT-1 weighted prevalence. We replicated the simulation 100 times to reduce sensitivity to sources of randomness. A key challenge here was to decide an appropriate benchmark against which we could compare accuracy of prevalence estimates, in survey rounds of varying sizes, with and without a wastewater model. In the absence of knowing the true prevalence in communities, we used a spatially-smoothed version of reported REACT-1 prevalence levels, thus reducing the likelihood of bias and misleading conclusions which could arise by benchmarking against either complete survey-based prevalence estimates or wastewater-model-based estimates. Briefly, the degree of spatial smoothing of LTLA-level prevalence observations within each round was determined by fitting of Kriging models to each of the 315 observations and the distances between LTLA centroids, thus accounting for the degree of spatial autocorrelation in prevalence levels within each survey round. Sensitivity of inferences to the choice of spatial smoothing method was appraised by also smoothing according to a smoothing kernel.

Wastewater-model-based estimates were appraised, relative to REACT-1 SARS-CoV-2 prevalence (unsmoothed unless stated otherwise), using metrics such as Mean Absolute Error (MAE), correlation with the response, and accuracy in detecting directional prevalence movements. As MAE is generally dependent on the prevalence levels of a specific round, we report the mean prevalence level to provide context for each attained MAE.

## Acknowledgements

The authors would like to thank several people who offered their valuable advice and expertise. We are grateful to Nicholas Steyn at the University of Oxford whose discussions provided important feedback for the analysis. Furthermore, we thank Matthew Wade, Till Hoffman, Guangquan Li, and Kathleen O’Reilly for sharing their wastewater expertise.

## Author Contributions

Conceptualization: C.A.D; C.M. Methodology: C.A.D; C.M; P.E; M.C.H. Data curation: C.M. Data visualisation: C.M. Writing original draft: C.M; C.A.D. All authors approved the final submitted draft.

## Data Availability

Access to REACT-1 individual-level data is restricted to protect participants’ anonymity. LTLA-level REACT-1 data can be obtained under data request. The wastewater concentration surveillance data from the EMHP are publicly available at https://www.gov.uk. The wastewater catchment areas for the United Kingdom are available at https://github.com/tillahoffmann/wastewater-catchment-areas.

