## Supplementary Materials for "Strategic use of SARS-CoV-2 wastewater concentration data could enhance, but not replace, high-resolution community prevalence survey programmes"

### Supplementary Material 1 Relationship Between SARS-CoV-2 RNA Wastewater Concentrations and Prevalence

Table SI 1: **Correlation analysis for wastewater concentrations and prevalence.** Spearman's correlation ($r$) is measured across each study period, where $n$ is the number of observations at each spatial resolution and a 95% CI is approximated using a paired bootstrap of 1000 replicates. Among LTLAs which report at least three wastewater concentration measurements across rounds 3 to 11 (24 July 2020 to 3 May 2021), the median LTLA-level correlation is 0.72, calculated as the median of each LTLA's individual correlation between wastewater concentrations and prevalence levels across the rounds. The corresponding median LTLA correlation between concentration-vaccination interaction and SARS-CoV-2 prevalence is 0.79 across rounds 12 to 19 (20 May 2021 to 31 March 2022).

| **Rounds** | **Variable** | **Spatial Resolution** | **n** | **r (95% CI)** |
|---|---|---|---|---|
| 3-11 | Concentration | LTLA | 1572 | 0.62 (0.59, 0.65) |
| 3-11 | Concentration | Regional | 81 | 0.83 (0.75, 0.89) |
| 3-11 | Concentration | National | 9 | 0.98 (0.82, 1.00) |
| 12-19 | Concentration-Vaccination Interaction | LTLA | 2461 | 0.71 (0.69, 0.73) |
| 12-19 | Concentration-Vaccination Interaction | Regional | 72 | 0.89 (0.79, 0.94) |
| 12-19 | Concentration-Vaccination Interaction | National | 8 | 0.93 (0.41, 1.00) |

### Supplementary Material 2 Wastewater-Based Model Covariates

Table SI 2: **Covariates used in wastewater-based models for estimating SARS-CoV-2 prevalence**. Unless otherwise stated, all covariates are at an LTLA level. Data originate from the REACT-1 study and the EMHP surveillance programme, aside from the LTLA population estimates (provided by the ONS) which are used as weights for regional and national predictors.

| **Variable** | **Units** | **Description** |
|---|---|---|
| Wastewater Concentration | Gene copies per litre (gc/l) | Estimated LTLA-level wastewater viral concentrations per round. Concentrations are obtained via the weighted contribution of each treatment plant's average (normalised) concentration for the round (described in Materials and Methods). |
| Neighbour-Averaged Concentration | Gene copies per litre (gc/l) | A spatial weights matrix (with row standardisation) is computed based on the neighbourhood structure of LTLAs, using queen contiguity criterion. The weights of this sparse matrix are multiplied by the LTLAs' concentrations for each round to yield a weighted average of the neighbouring areas' concentrations. |
| Vaccination-Log Concentration Interaction | Gene copies per litre (gc/l) | (Fully Vaccinated Proportion) $\times$ (Estimated Concentration). Fully vaccinated proportions are the proportion of the LTLA population estimated (by the REACT-1 study) to have received 2 or more vaccination doses, whilst concentrations are the estimated LTLA-level concentration. |
| Regional Average Concentration | Gene copies per litre (gc/l) | Average of a region's wastewater concentrations for a round, weighted by the underlying LTLA populations (for whom measurements were available in that round). |
| National Average Concentration | Gene copies per litre (gc/l) | National average of wastewater concentrations for a round, weighted by the underlying LTLA populations (for whom measurements were available in that round). |
| Concentration Difference | Gene copies per litre (gc/l) | The change in an LTLA's wastewater concentration from one round to the next. |
| National Concentration Difference | Gene copies per litre (gc/l) | The change in the national average wastewater concentration from one round to the next. |
| Regional Prevalence per Log Concentration | % per gc/l | For each LTLA, prevalence per log concentration is calculated, and the corresponding regional average is the weighted average of the LTLA-level values for that round (where the weights are the LTLA populations). The regional average across the training period is used as the predictor for the testing set. Region-level prevalence per log concentration is used due to the noisiness of the LTLA-level estimates. Prevalence per log concentration can be interpreted as a proxy for population-level faecal shedding dynamics. Hence, greater values of the prevalence-to-wastewater variable implies a reduced level of faecal shedding per positive individual. |

### Supplementary Material 3 Out-of-Sample Wastewater-Model-Based Estimates of Prevalence

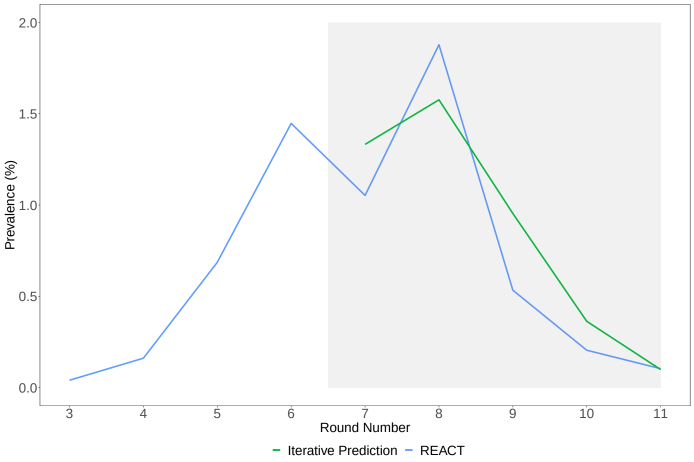


Figure SI 1: **Iteratively-updated model-based national prevalence estimates in rounds 7-11 (13 November 2020 to 3 May 2021)**. Visualisation of national-level gradient boosting model predictions (green) alongside REACT-1 prevalence (blue), for five rounds of REACT-1 study.

Table SI 3: **Regional wastewater-model-based estimates for REACT-1 rounds 9-11 (4 February 2021 to 3 May 2021)**. Summary of accuracy metrics for regional estimates, drawing comparison across out-of-sample estimates for individual rounds for a model i) which is trained iteratively and ii) which makes multi-step estimates without any model updating/calibration. Accuracy measures maintain the same interpretation as Table SI 4, albeit now at a regional level. Average Prevalence is 0.31%, calculated as the mean regional prevalence across rounds 9-11. * The iterative training method involves updating the wastewater model using data from the additional training round which precedes the testing round.

| **Training Method** | **Testing Rounds** | **Training Rounds** | **n** | **MAE** | **r** | **Change Detection (95% CI)** |
|---|---|---|---|---|---|---|
| Iterative | 9-11 | 3-10* | 27 | 0.20% | 0.93 | 85.2% (66.3%, 95.8%) |
| Multi-Step | 9-11 | 3-8 | 27 | 0.26% | 0.84 | 70.4% (49.9%, 86.3%) |

Table SI 4: **Out-of-sample LTLA-level predictive performance for REACT-1 rounds 7-11 (13 November 2020 to 3 May 2021).** Out-of-sample estimates are based on a wastewater-based gradient boosting model trained iteratively, including for the regional (*) wastewater-based estimates. *n* is the total number of LTLAs in an individual testing round and in the regional (**) case, the total number of regional observations from rounds 7-11. *MAE* is the Mean Absolute Error between model-based estimates and REACT-1 prevalence. *r* represents the Pearson's correlation between the prevalence and our estimates. *Top 25 Common* is the number of LTLAs common to the highest predicted prevalence levels and REACT-1 prevalence levels. *Change Detection* indicates the mean directional accuracy, whilst the corresponding 95% CI is attained by the so-called Clopper-Pearson method, otherwise known as the Exact Confidence Interval [32]. *Mean Prevalence* is the mean average of LTLA-level REACT-1 prevalence estimates (for LTLAs in the wastewater surveillance programme), and is cited as a guide for appraisal of MAE within rounds. We provide the individual round analysis of *regional* model-based estimates performance in Table SI 9

| **Test Round** | **Training Rounds** | **n** | **MAE** | **r** | **Top 25 Common** | **Change Detection (95% CI)** | **Mean Prevalence** |
|---|---|---|---|---|---|---|---|
| 7 | 3-6 | 146 | 0.7% | 0.08 | 4 | 66.4% (58.2%, 74.0%) | 1.0% |
| 8 | 3-7 | 146 | 1.0% | 0.07 | 4 | 69.9% (61.7%, 77.2%) | 1.8% |
| 9 | 3-8 | 233 | 0.5% | 0.27 | 8 | 76.0% (70.0%, 81.3%) | 0.5% |
| 10 | 3-9 | 299 | 0.3% | 0.23 | 4 | 69.2% (63.7%, 74.4%) | 0.2% |
| 11 | 3-10 | 301 | 0.2% | 0.08 | 1 | 57.9% (52.0%, 63.5%) | 0.1% |
| Regional: 7-11 | 3-10* | 45 ** | 0.3% | 0.76 | - | 86.7% (76.7%, 96.6%) | 0.7% |

### Supplementary Material 4 Complementary Use of WBE

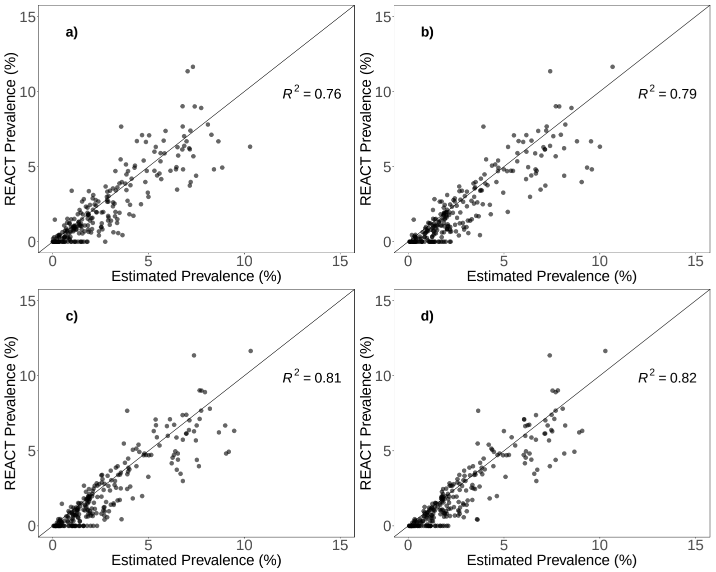


Figure SI 2: **Wastewater-based prevalence estimates using varying training set sizes.** From plots a) to d), across rounds 12 to 19 (from 20 May 2021 to 31 March 2022), training set sizes are 40%, 60%, 80%, and 90% respectively per each round's total number of LTLAs. The plot above is for a randomly-selected fold amongst the 50 folds considered in our replicated procedure for training-testing split proportions, and model-based prevalence estimates of a fixed test set improve marginally as more training observations are used to calibrate the model. Nevertheless, with just 40% survey coverage, wastewater-model-based estimates remain largely representative of underlying prevalence.

Table SI 5: **Comparison of wastewater-based prevalence estimates in the *early* period of rounds 3 to 11 (24 July 2020 to 3 May 2021) using varying training test set sizes**. Wastewater-based gradient boosting models are trained using 40%-90% of each round's observations and a fixed 10% of observations per round are used for test set prevalence estimation. The average prevalence across the 50 folds of test sets is 0.54%.

| Training-Testing | MAE | r | Change Detection |
|---|---|---|---|
| 40% - 10% | 0.34 % | 0.74 | 74.57% |
| 60% - 10% | 0.33 % | 0.74 | 75.14% |
| 80% - 10% | 0.32 % | 0.76 | 76.30% |
| 90% - 10% | 0.32 % | 0.75 | 74.57% |

### Supplementary Material 5 Population-level faecal shedding

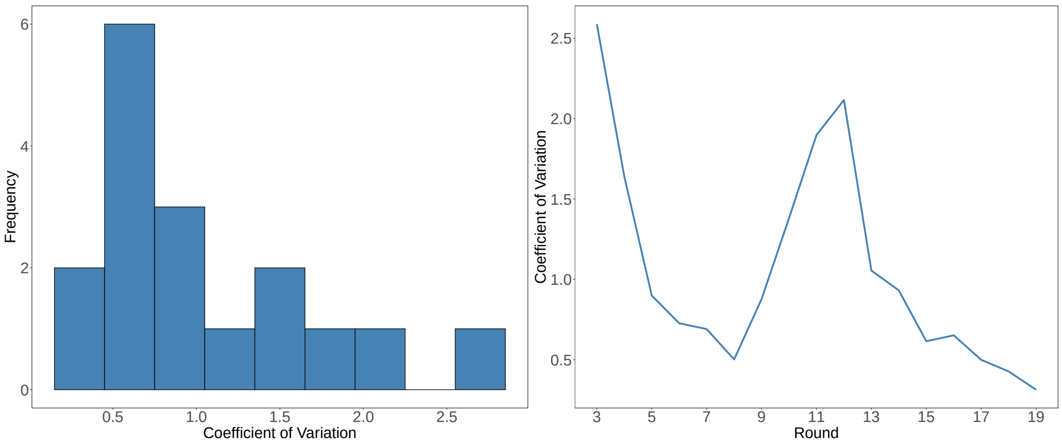


Figure SI 3: **Spatial variability of faecal shedding by round.** Visualisations demonstrate the spatial consistency of the prevalence-to-wastewater relationship, particularly during enhanced wastewater surveillance in the later rounds of the REACT study. The histogram (left) of the coefficient of variation (CV) values demonstrate that extreme large values of the measure of dispersion are not common, whilst the plot over time (right) captures that the larger values occur during early rounds with low wastewater surveillance coverage and/or rounds of low prevalence.

#### Supplementary Material 6 Geospatial Population Estimates (GPEs)

The following section provides further detail regarding the derivation and uncertainty of the GPEs used for our geospatial framework and associated analyses.

Let $a_{ij}$ be the intersection area between the catchment area of STW $i$ and LSOA $j$. Then, the LSOA area serviced by any of the STWs $i$ is:

$$A_j = \bigcup_i a_{ij}$$

Let $p_j$ be the Office for National Statistics (ONS) 2019 mid-year population estimate for LSOA $j$. Our geospatial population estimate (GPE) for the intersection area between the catchment area of STW $i$ and the area of LSOA $j$ is

$$\widehat{g_{ij}} = \frac{a_{ij}}{A_j} p_j$$

Thus, for LSOA $j$, our estimate of the LSOA population serviced is:

$$\widehat{p_j} = \sum_i \widehat{g_{ij}}$$

Summing over all LSOA geographies within an LTLA $k$, we attain our GPE $\widehat{P_k}$ for each LTLA:

$$\widehat{P_k} = \sum_{j=1}^{J} \widehat{p_j}$$

The GPE for each LTLA enables our mapping of concentrations from an STW level to an LTLA level.

Our approach of using GPEs represents a predictive approach for aligning WBE with the geographies commonly used by public health authorities [28]. Consistent with the reported EMHP population coverage estimates [14], we estimate that the wastewater programme had an estimated 74% nationwide testing coverage by its conclusion in March 2022, with a median percentage (of population) sampled for LTLAs of 76%.

The developed approach could, in theory, be applicable to further population-level wastewater monitoring and analyses which require alignment of wastewater catchments to geographies used by public health authorities and community studies. Nevertheless, limitations exist with our geospatial approach. First, the GPEs do not account for the presence of transient and non-resident LTLA populations, which could impact on the wastewater concentrations measured. The transience issue is likely to be more pronounced in the late rounds of the REACT-1 analysis, where community lockdowns and other non-pharmaceutical interventions are relaxed. Second, we recognise the uncertainty surrounding the mid-year population estimates reported by the ONS for LSOAs, which are made for small geographic areas.

Furthermore, potential limitations may yield unrepresentative population estimates for urban centres and/or for geographies which are influenced by time-varying factors such as transient/commuting populations or industrial and agricultural discharges. In theory, such factors are controlled via our flow-normalised wastewater concentrations yet cannot be taken into account when we weight the contribution of each STW to an LTLA based on time-invariant population estimates). The issue of time-varying relationships is likely to depend upon the type and stringency level of simultaneously active non-pharmaceutical interventions. Further challenges imposed by the EMHP wastewater surveillance programme include the locations of the 302 STWs. Whilst the sampled treatment plants were selected to maximise nationwide coverage and representativeness across England, our GPEs (for each LTLA) indicate a highly positively skewed distribution for the estimated proportions of individual LTLA populations that are sampled within the EMHP programme (Figure SI 4). The skewness in the distribution of estimated proportions of populations sampled is is a likely consequence of the usage of intersection areas in our GPEs. We estimate the median LTLA-level proportion of population sampled to be approximately 76%, yet Horesham LTLA, for instance, has an estimated testing coverage of only 2.9% based on the GPE, consistent with the EMHP testing coverage estimate for Horesham. These few outliers can potentially impact on how representative our wastewater concentrations are when mapped to LTLAs with such low testing coverage, and hence, when relating our wastewater data to LTLA-level survey-based prevalence estimates.

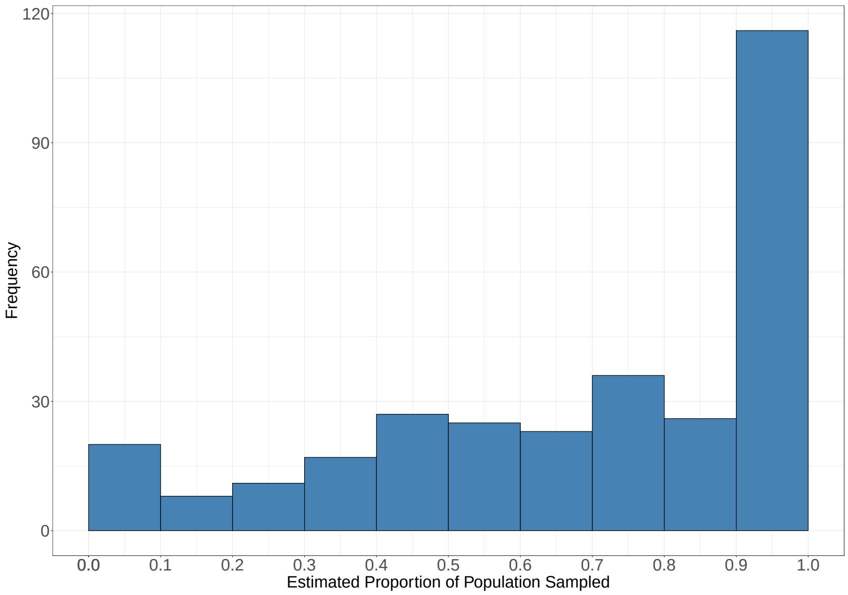


Figure SI 4: **LTLA-level wastewater sampling coverage estimates.** The estimated proportions of LTLA populations that are covered by the EMHP wastewater surveillance programme. The proportions are attained by dividing the LTLA-level GPE by the corresponding ONS LTLA population approximation.

### Supplementary Material 7 Wastewater Sources of Uncertainty

Sources of wastewater measurement uncertainty include the sample volume being too low to enable adequate analysis, temperature and time-induced decay, inherent variability of wastewater (possibly due to dilution effects of precipitation), and usage of the Theoretical Limit of Detection (TLoD), below which the concentration cannot be reliably measured. The EMHP surveillance programme aimed to address several of these measurement uncertainties via the adjustment/normalisation of concentrations to account for flow, and by sampling mid-stream during peak load times. Similarly, 3 to 4 samples were taken weekly (at each STW) due to the variability and presence of outliers in detected wastewater signals [14, 22].

### Supplementary Material 8 Sensitivity of Geospatial Mapping Approach

Table SI 6: **Sensitivity analysis for geospatial mapping**. Table depicts a diminished relationship between wastewater concentrations and SARS-CoV-2 prevalence, when concentrations from rounds of the REACT-1 are shifted by a lead times of up to six days. Overall correlation measures the correlation between LTLA-level estimated wastewater concentrations and SARS-CoV-2 prevalence. Mean LTLA concentration correlation is the mean of all the LTLA's individual correlations between estimated wastewater concentrations and SARS-CoV-2, whilst the mean LTLA interaction correlation (applicable for rounds 12 to 19) is the average of the LTLA's individual correlation between our proposed vaccination-log concentration interaction and SARS-CoV-2 prevalence.

| Lead Time (Days) | Overall Correlation | Mean LTLA Concentration Correlation | Mean LTLA Interaction Concentration Correlation |
|---|---|---|---|
| 0 | 0.35 | 0.39 | 0.75 |
| 1 | 0.32 | 0.33 | 0.75 |
| 2 | 0.29 | 0.31 | 0.75 |
| 3 | 0.28 | 0.30 | 0.75 |
| 4 | 0.28 | 0.25 | 0.74 |
| 5 | 0.27 | 0.25 | 0.74 |
| 6 | 0.19 | 0.16 | 0.66 |

### Supplementary Material 9 Bayesian Hierarchical Modelling

To ensure robustness of inferences and extend our analysis beyond a discriminative model, within the Bayesian modelling paradigm, we fitted a hierarchical model, which is a generative model type which enables regularisation and spatial effects. Our hierarchical model is fit to training periods using Stan v2.31, which employs Hamiltonian Monte Carlo sampling, a variant of Markov Chain Monte Carlo (MCMC) [33].

Let $y_{it}$ be the REACT-1 weighted prevalence estimate for LTLA $i$ at time/round $t$. Then, our varying effects Bayesian hierarchical model is as follows:

$$
\begin{aligned}
y_{it} &\sim \text{Normal}(\mu_{it}, \sigma^2) \\
\mu_{it} &= \beta_0 + \beta_{LTLA[i]} * C_{it} + \beta_{R[i]} * C_{it} + \beta_{NN[i]} * NC_{it} \\
\beta_0 &\sim \text{Normal}(0, 5) \\
\beta_{LTLA} &\sim \text{Normal}(0, \sigma^2_{LTLA}) \\
\sigma_{LTLA} &\sim \text{InvGamma}(5, 5) \\
\beta_R &\sim \text{Normal}(0, \sigma^2_R) \\
\sigma_R &\sim \text{InvGamma}(5, 5) \\
\beta_{NN} &\sim \text{Normal}(0, \sigma_{NN}) \\
\sigma_{NN} &\sim \text{InvGamma}(5, 5) \\
\sigma &\sim \text{Half-Cauchy}(0, 1)
\end{aligned}
$$

where $C_{it}$ denotes the concentration of LTLA $i$ at time/round $t$, and $NC_{it}$ denotes the weighted average concentration for the neighbours of LTLA $i$ at time/round $t$. $\mu$ and $\sigma$ represent the mean and standard deviation of the assumed Normal distribution. $\beta_0$ is the baseline prevalence. $\beta_{LTLA}$, $\beta_R$, and $\beta_{NN}$, are parameter vectors corresponding to the LTLA-specific, region-specific, and nearest-neighbour specific effects of concentration on the response of prevalence. $\sigma_{LTLA}$, $\sigma_R$, and $\sigma_{NN}$ represent the corresponding standard deviations. Above, we employ an index-variable approach within the linear predictor. For example $\beta_{R[i]}$ maps the LTLA $i$ to its corresponding parameter within the region-effect concentration vector. The intuition for spatially-varying effects is that in-sewer network characteristics, treatment plant-level, and spatial correlations between geographies could influence similarities and differences of both concentrations and prevalence levels. For the *late* period of the REACT-1 analysis, the vaccination-log concentration interaction (described in Table SI 2) takes the place of each of the unadjusted wastewater concentrations.

Throughout the analysis, all covariates are standardised such that they take values in $[-1, 1]$. Standardisation facilitates the above usage of conventional, weakly informative prior distributions throughout our model fitting, and assists in ensuring efficient HMC sampling.

Similar to the gradient boosting setup, the model was trained using several calibration rounds from REACT-1, and its predictive performance was estimated out-of-sample using one or more rounds of REACT-1. Our model selection procedure involved consideration of the scientific model's structure, posterior predictive checks, model convergence diagnostics, and estimated out-of-sample predictive accuracy (via the ELPD, expected log pointwise predictive density, from the `loo` package).

In terms of comparing predictive performance to our gradient boosting model, we generate posterior predictions of the response $y_{it}$. In particular, we use the posterior samples for each observation, and we adopt a conventional approach of using the posterior median of these samples as our best estimate (for each observation). We quantify the uncertainty in our inferences by deriving credible intervals (by taking specified upper and lower quantiles of these samples). Subsequently, metrics such as MAE, for example, are computed as the average absolute difference between the observed prevalence levels and the posterior median estimates.

Throughout both study periods, our best-performing Bayesian hierarchical model did not provide additional predictive accuracy in terms of out-of-sample predictive accuracy. Superior accuracy of the gradient boosting model may be a consequence of the highly flexible and predictive nature of the modern gradient boosting algorithm (of `xgboost`) which readily handles time-varying non-linearites. Nevertheless, the additional modelling investigation enabled a further robustness check of the inferences drawn regarding the reliability of wastewater-based modelling.

### Supplementary Material 10 Vaccination-Log Concentration Interaction Variable

In terms of the statistical controls in our analysis, there exist obvious limitations with our proposed usage of a simple vaccination-concentration interaction. Aside from the potential differential impacts of vaccination on distinct demographies, the proposed interaction variable would only remain (possibly) valid for a restricted time period due to impacts of waning vaccine-induced immunity. We further do not account for the highly complex nature of naturally-acquired immunity. The interaction variable's usage is additionally challenged by the disparate impact of different variants (and sub-variants) on the likelihood of SARS-CoV-2 reinfection. Importantly, we do not draw *any causal conclusions* regarding the impact of vaccination on faecal shedding due to the potential presence of confounding. Indeed, we emphasise, that strong temporal correlation is not necessarily indicative of a direct (or causal) relationship with prevalence, as the directional association may be an artefact of numerous epidemic conditions. Complex, concurrent epidemic characteristics include a monontonically increasing vaccination proportion, similarly rising prevalence levels, waning immunity (either vaccine-induced or naturally-acquired), reduced immunity to particular new variants, and other possibly unobserved confounders.

### Supplementary Material 11 Acronyms

Table SI 7: **Acronyms used throughout this paper.**

| **Variable** | **Meaning** | **Details** |
|---|---|---|
| 95% CrI | 95% Credible Interval | An unobserved parameter lies in this interval with a specified 95% probability. |
| EMHP | Environmental Monitoring for Health Protection | The EMHP wastewater surveillance programme tested untreated sewage across England for fragments of SARS-CoV-2. The objectives of the programme were to monitor wastewater viral concentrations of SARS-CoV-2 RNA, variants of concern (VOC), and variants under investigation (VUI). |
| gc/l | Gene copies per litre (gc/l) | The reported measuresments of wastewater concentrations obtained via RT-qPCR and flow normalisation. |
| GPE | Geospatial population estimate | Our population estimates for geographies based on a combination of intersection (spatial) areas and reported population estimates for small geographies. |
| LSOA | Lower Layer Super Output Area | Small regional geographies in England which combine to form an LTLA. |
| LTLA | Lower Tier Local Authority | LTLA-level wastewater measurements per round. These are obtained via the weighted contribution of each treatment plant's average concentration for the round (described in Materials and Methods). |
| MAE | Mean Absolute Error | The mean average of the absolute difference between the model-based estimates and the REACT-1 prevalence levels. |
| MCMC | Markov Chain Monte Carlo | A family of sampling algorithms which employs the theory of Markov Chains to sample a random variable. |
| NPI | Non-pharmaceutical intervention | In the context of the COVID-19 pandemic, these are public health measures, excluding medication-based measures, taken to control transmission of the SARS-CoV-2. |
| ONS | Office for National Statistics | An independent producer of national statistics across the UK. |
| REACT-1 | Real-time Assessment of Community Transmission | The REACT-1 programme was initiated in May 2020 with an objective of tracking the spread of SARS-CoV-2 across communities in England. Across 19 distinct rounds of cross-sectional surveys, random samples of the English population (over 5 years of age) were taken. Rounds lasted between 15 and 31 days, and the programme concluded in March 2022. |
| RNA | Ribonucleic acid | RNA is a nucleic acid present in all living cells. |
| RT-PCR | Gene copies per litre (gc/l) | LTLA-level wastewater measurements per round. These are obtained via the weighted contribution of each treatment plant's average concentration for the round (described in Matrerials and Methods). |
| RT-qPCR | Reverse Transcriptase Polymerase Chain Reaction (RT-qPCR) | The quantification method is described in [7]. Briefly, RT-qPCR combines reverse transcription and quantitative PCR, with the aim of reducing inhibition in RNA. |
| STW | Sewage treatment works | STWs are treatment plants which typically service extensive urban areas (like towns and cities). |
| TLoD | Theoretical limit of detection | The wastewater concentration (160 gc/l) below which the EMHP estimate that concentration cannot be reliably estimated. |
| UKHSA | United Kingdom Health Security Agency | A nationwide organisation in the United Kingdom which assumes responsibility for public health protection. |
| VOC | Variants of Concern | SARS-CoV-2 variants that were highlighted by the World Health Organisation (WHO) to be particularly dangerous in terms of increased transmissibility. |
| VUI | Variants under investigation | SARS-CoV-2 variants were being tracked by the EMHP wastewater surveillance programme. |
| WBE | Wastewater-based epidemiology | WBE involves collection of urine and stool samples from sewage treatment works (STWs). By subsequently incorporating factors such as daily flow rates, human excretion rates, and STW catchment population sizes, per-capita consumption, use, or exposure can be obtained. |
| WHO | World Health Organisation | An agency of the United Nations which aims to improve international public health. |

### Supplementary Material 12 Timeline of REACT-1 and Estimated Number of LTLAs in EMHP Programme

Table SI 8: Dates (DD/MM/YYYY) for individual rounds of the REACT-1 study and the corresponding estimated number of LTLAs which were mapped to STWs which reported measurements within the corresponding time intervals. Our overall studied period, where the EMHP wastewater surveillance programme coincides with the REACT-1 study, covers from rounds 3 to 19 (from 24 July 2020 to 31 March 2022.

| Round Number | Round Start Date | Round End Date | Number of LTLAs |
|---|---|---|---|
| 1 | 01/05/2020 | 01/06/2020 | - |
| 2 | 19/06/2020 | 07/07/2020 | - |
| 3 | 24/07/2020 | 11/08/2020 | 145 |
| 4 | 20/08/2020 | 08/09/2020 | 145 |
| 5 | 18/09/2020 | 05/10/2020 | 146 |
| 6 | 16/10/2020 | 02/11/2020 | 146 |
| 7 | 13/11/2020 | 03/12/2020 | 146 |
| 8 | 06/01/2021 | 22/01/2021 | 146 |
| 9 | 04/02/2021 | 23/02/2021 | 233 |
| 10 | 11/03/2021 | 30/03/2021 | 299 |
| 11 | 15/04/2021 | 03/05/2021 | 301 |
| 12 | 20/05/2021 | 07/06/2021 | 303 |
| 13 | 24/06/2021 | 12/07/2021 | 306 |
| 14 | 09/09/2021 | 27/09/2021 | 307 |
| 15 | 19/10/2021 | 05/11/2021 | 309 |
| 16 | 23/11/2021 | 14/12/2021 | 309 |
| 17 | 05/01/2022 | 20/01/2022 | 309 |
| 18 | 08/02/2022 | 01/03/2022 | 309 |
| 19 | 08/03/2022 | 31/03/2022 | 309 |

### Supplementary Material 13 Wastewater Variant Detections

From a public health perspective and for our analysis, the ability to track variants is important to understand the relationship between wastewater concentrations and our community prevalence estimates. Specifically, the EMHP programme raised the conjecture of an Omicron effect which results in lower viral faceal shedding and substantially alters the relationship between wastewater signals and clinical measures [14]. The variant detections were attained via genomic sequencing of wastewater samples from both STWs and sewer network sites across England, and detections were reported as either a confirmed or a possible status. More recently, the importance of accounting for the predominant variant was demonstrated by research of clinical cases showing that Omicron infections yielded the lowest community-level SARS-CoV-2 waste shedding rates, compared to the early/parental SARS-CoV-2 and Delta variant [20]. Within our analysis, we have documented the apparent reduced population-level faecal shedding induced by the onset of the Omicron BA.1 and BA.2 sub-variants.

### Supplementary Material 14 Further Results

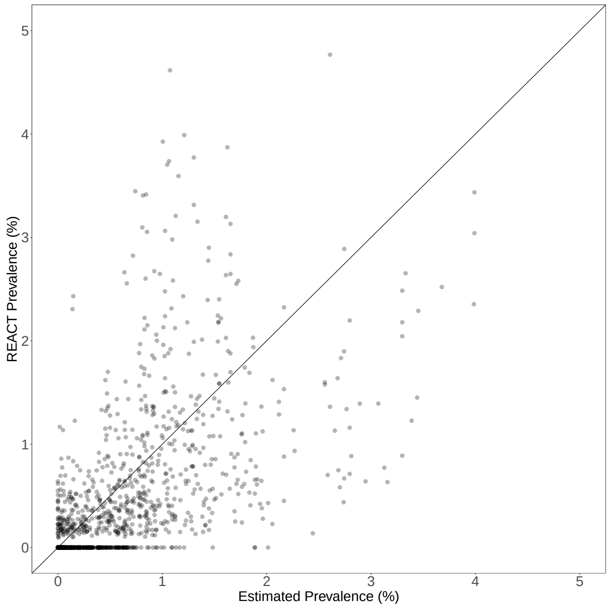


Figure SI 5: **Iteratively-updated wastewater-model-based prevalence estimates for rounds 7 to 11 (13 November 2020 to 3 May 2021).** Visualisation depicts REACT-1 prevalence versus wastewater-model-based estimates for the corresponding round.

Table SI 9: **Regional out-of-sample predictive performance by round**. Summary of regional out-of-sample wastewater-based predictive performance by round for REACT-1 rounds 7 to 11 (13 November 2020 to 3 May 2021) and rounds 15 to 19 (19 October 2021 to 31 March 2022), where out-of-sample wastewater-based estimates were obtained based on a gradient boosting model trained iteratively at an LTLA level. Measures of accuracy have the same interpretation as Table SI 4, albeit now at a regional level. Average regional prevalence is provided as a reference for accuracy measures.

| Testing Round | Training Rounds | n | MAE | Change Detection (95% CI) | Average Prevalence |
|---|---|---|---|---|---|
| 7 | 3-6 | 9 | 0.37% | 66.7% (29.9%, 92.5%) | 1.0% |
| 8 | 3-7 | 9 | 0.70% | 88.9% (51.8%, 99.7%) | 1.6% |
| 9 | 3-8 | 9 | 0.37% | 88.9% (51.8%, 99.7%) | 0.5% |
| 10 | 3-9 | 9 | 0.17% | 88.9% (51.8%, 99.7%) | 0.2% |
| 11 | 3-10 | 9 | 0.05% | 77.8% (40.0%, 97.2%) | 0.1% |
| 15 | 12-14 | 9 | 0.47% | 100.0% (66.4%, 100.0%) | 1.6% |
| 16 | 12-15 | 9 | 0.50% | 55.6% (21.2%, 86.3%) | 1.4% |
| 17 | 12-16 | 9 | 2.88% | 100.0% (66.4%, 100.0%) | 4.7% |
| 18 | 12-17 | 9 | 1.21% | 77.8% (40.0%, 97.2%) | 2.8% |
| 19 | 12-18 | 9 | 1.63% | 100.0% (66.4%, 100.0%) | 6.3% |

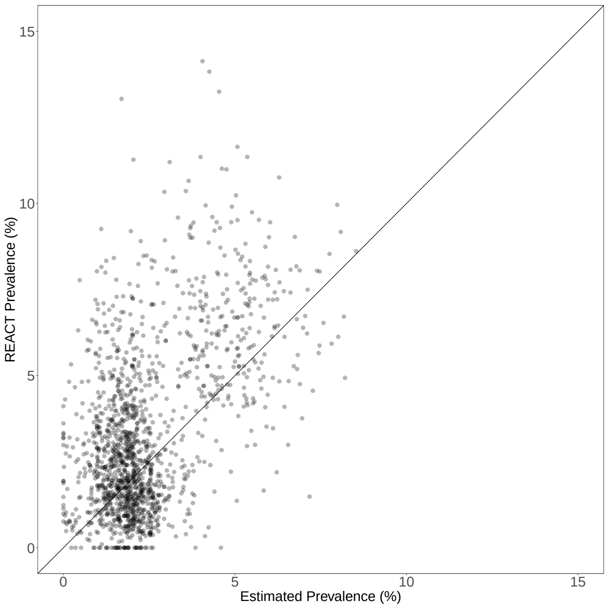


Figure SI 6: **Iteratively-updated wastewater-model-based prevalence estimates for rounds 15 to 19 (19 October 2021 to 31 March 2022).** Visualisation depicts REACT-1 prevalence versus wastewater-model-based estimates for the corresponding round.

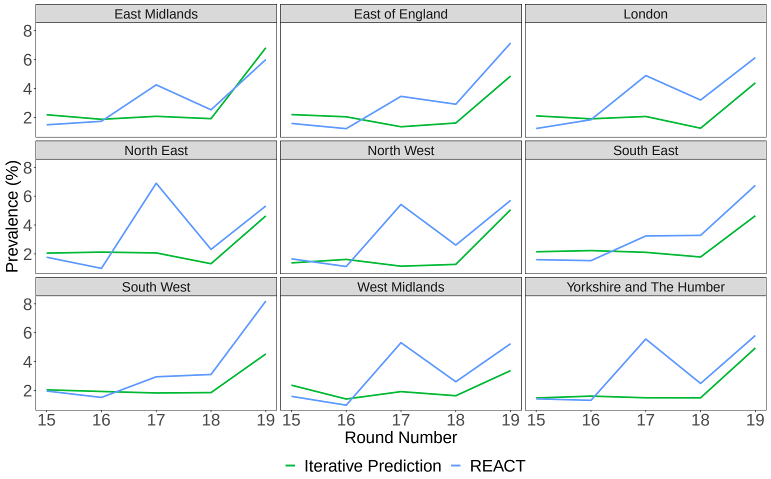


Figure SI 7: **Iteratively-trained wastewater model's prevalence estimates in rounds 15 to 19 (19 October 2021 to 31 March 2022).** Gradient Boosting model's regional estimates are shown (green), alongside REACT-1 prevalence (blue). All out-of-sample estimates are made iteratively for individual (single round) test sets. The key insights are i) the Omicron BA.1 peak (round 17) is undetected by the calibrated wastewater model and its concentrations, and ii) re-calibration during the Omicron wave enables identification of rises in regional prevalence in the Omicron BA.2 peak (Round 19).

Table SI 10: **Out-of-sample predictive performance for REACT-1 rounds 15-19 (19 October 2021 to 31 March 2022),**. LTLA-level predictions are made based on a gradient boosting model trained iteratively. All accuracy measures are defined as reported in Table SI 4.

| Testing Round | Training Rounds | n | MAE | r | Top 25 Common | Change Detection (95% CI) | Average Prevalence |
|---|---|---|---|---|---|---|---|
| 15 | 12-14 | 309 | 0.9% | 0.02 | 2 | 79.0% (74.3%, 83.7%) | 1.6% |
| 16 | 12-15 | 309 | 1.0% | 0.09 | 0 | 68.9% (62.8%, 73.4%) | 1.4% |
| 17 | 12-16 | 309 | 2.5% | 0.04 | 3 | 68.6% (63.1%, 73.7%) | 4.2% |
| 18 | 12-17 | 309 | 1.5% | 0.03 | 2 | 75.7% (70.6%, 80.4%) | 2.8% |
| 19 | 12-18 | 309 | 2.2% | -0.02 | 1 | 86.0% (81.7%, 89.7%) | 6.5% |
